# Phylotyping, Pathotyping and Phenotypic Characteristics of *Escherichia coli* Isolated from Various Street Foods in Bangladesh

**DOI:** 10.1101/2022.02.07.22270615

**Authors:** Anindita Bhowmik, Sharmistha Goswami, Ahmad Salman Sirajee, Sunjukta Ahsan

## Abstract

The burden of street food contamination remains significantly higher among developing countries, making its consumers’ health compromised. This study aimed at identifying *Escherichia coli* in local street foods, track their sources and characterize their virulence properties, antimicrobial susceptibility patterns, and incidence of recent fecal contamination. Twelve *E. coli* isolates isolated from 28 types of street foods sold at different locations of Dhaka metropolitan city were confirmed by both culture-based and molecular methods. Phylogroup B1 of environment origin was the most predominant (58%, n=12) among the isolates, followed by commensal phylogroup A (16%), phylogroup C (8%), D (8%), and E (8%). Alarmingly, 8% of the isolates were confirmed to be Enteropathogenic *E. coli* (EPEC), harboring *hly*A gene, and 41% were due to recent fecal contamination cases confirmed by the presence of the *eae* gene. Isolates showed the highest resistance to ampicillin, followed by chloramphenicol. However, resistance could not be correlated to the presence of class 1 integron as isolates sensitive to these antibiotics also harbored this mobile genetic element. The findings of this study demonstrated the presence of antibiotic-resistant, potentially pathogenic *E. coli* in street food which emphasizes the health hazard associated with consumption of such foods.

## Introduction

“Street food” refers to a wide variety of foods and beverages that are usually prepared or sold by vendors in streets or any public places for direct human ingestion immediately or later. These foods are generally low-priced, tasty, varied in types, and readily available (Sabuj et al., 2018). Street foods like shingara, samosa, vegetable roll, chicken roll, burger, sandwich, pakora, puri, chop, jhalmuri, chanachur, fuchka, chotpoti, cake, and patties are easy to prepare and serve. These foods have earned popularity among many low- and middle-income countries like Bangladesh (Rahman et al., 2014). Despite numerous benefits delivered by street foods, they have been reported to cause severe safety and health distresses among customers and food handlers. Inadequate food safety knowledge and proper personal hygiene practices along with lack of public awareness are responsible for increasing foodborne disease burden in developing countries (Birgen et al., 2020). Contaminated street food has been related to foodborne illness and foodborne outbreaks equally (Hassan et al., 2016). Therefore, foodborne infections have become a rising public health concern and an alarming issue worldwide, which cause up to 2 million deaths in unindustrialized countries each year (Rahman et al., 2017). It has accelerated the increasing antibiotic resistance and thus made available empirical treatment options limited. Worldwide, a statistical study declared that foodborne diseases caused by the catering industries are 70% greater than the food poisoning caused by another sector (Hassan et al., 2017). Around 30 million people get infected by foodborne diseases every year due to the fast-food ingredients in Bangladesh. It has become a source of many pathogens like *Escherichia coli, Staphylococcus aureus, Enterobacter* spp, *Salmonella enterica, Shigella* spp, and *Listeria monocytogenes*, although most of the constituents used in such foods have nutritive values (Sabuj et al., 2018). However, *Escherichia coli* receives more attention among all foodborne pathogens as it is a facultative anaerobic bacterium and is considered the most commonly encountered pathogen in the *Enterobacteriaceae* family. In addition, *E. coli* acts as a vital food hygiene indicator since its presence reflects the possibility of contamination by other enteric pathogens (Frisca et al., 2007; Biswas et al., 2010; Rahman et al., 2017).

Culture and molecular methods can help in the identification of *E coli*. In this regard, two *E. coli* specific genetic markers, *usp*A and *uid*A, encoding universal stress protein and β-glucuronidase enzyme, respectively, can be used for confirmation (Godambe et al., 2017). On the other hand, Amplified Ribosomal DNA Restriction Analysis (ARDRA) is considered a suitable and time-consuming way to provide correct subtyping results for this microorganism (Diijkshoorn et al., 1998; Jawad et al., 1998; Shin et al., 2004).

*E. coli* being a normal flora, turned out to be pathogenic due to the environment. Rampant antibiotic consumption at any clinical onset has made the situation even worse without any professional insight. Therefore multidrug-resistant *E. coli* has become a public health concern nowadays as this microbe can become an opportunistic pathogen. Its pathogenic mechanism includes adhesion and binding to the small intestine’s mucosal receptor (Mousa and Shama, 2020). One of the virulence genes of *E. coli, hly*A, which encodes Hemolysin A can create pores in the cell membranes of host cells and result in cell lysis (Sarowska et al., 2019).

Previous research on *E. coli* phylotypes shows that the contamination flora of meat and meat products are typically comprised of phylogenetic groups A, B, and D (Sunde et al., 2015). Clermont *et al*. revealed eight different phylogroups of *E. coli*, with seven belonging to *E. coli sensu stricto* (A, B1, B2, C, D, E, F) and one corresponding to *Escherichia* clade I (Clermont et al., 2013). Phylogroup A characterizes commensal *E. coli* strains (Picard et al., 1999), and phylogroup B1 represents strains that can persist in the environment (Walk et al., 2007). Phylogroups B2 and D include only virulent *E. coli* strains (Carlos et al., 2010). Phylogroup E is a new phylogroup that is unassigned. Phylogroup F is considered a sister group of phylogroup B2, and phylogroup C contains closely related strains but distinct from phylogroup B1 (Clermont et al., 2013). Some strains hold properties of both phylogroups F and B2 and are categorized as phylogroup G (Clermont et al., 2019).

The objectives of this study include isolation and identification of *E. coli* from street foods, determining their origin, profiling drug resistance patterns, and investigating the presence of mobile genetic elements like class 1 integrons and plasmids.

## Materials And Methods

### Sample collection

This research was carried out on street foods collected from different Dhaka metropolitan areas between March 2017 and May 2017. A total of 60 fast-food samples comprising 3 categories (Table 1) were collected from different roadside carts.

**Table 1.**
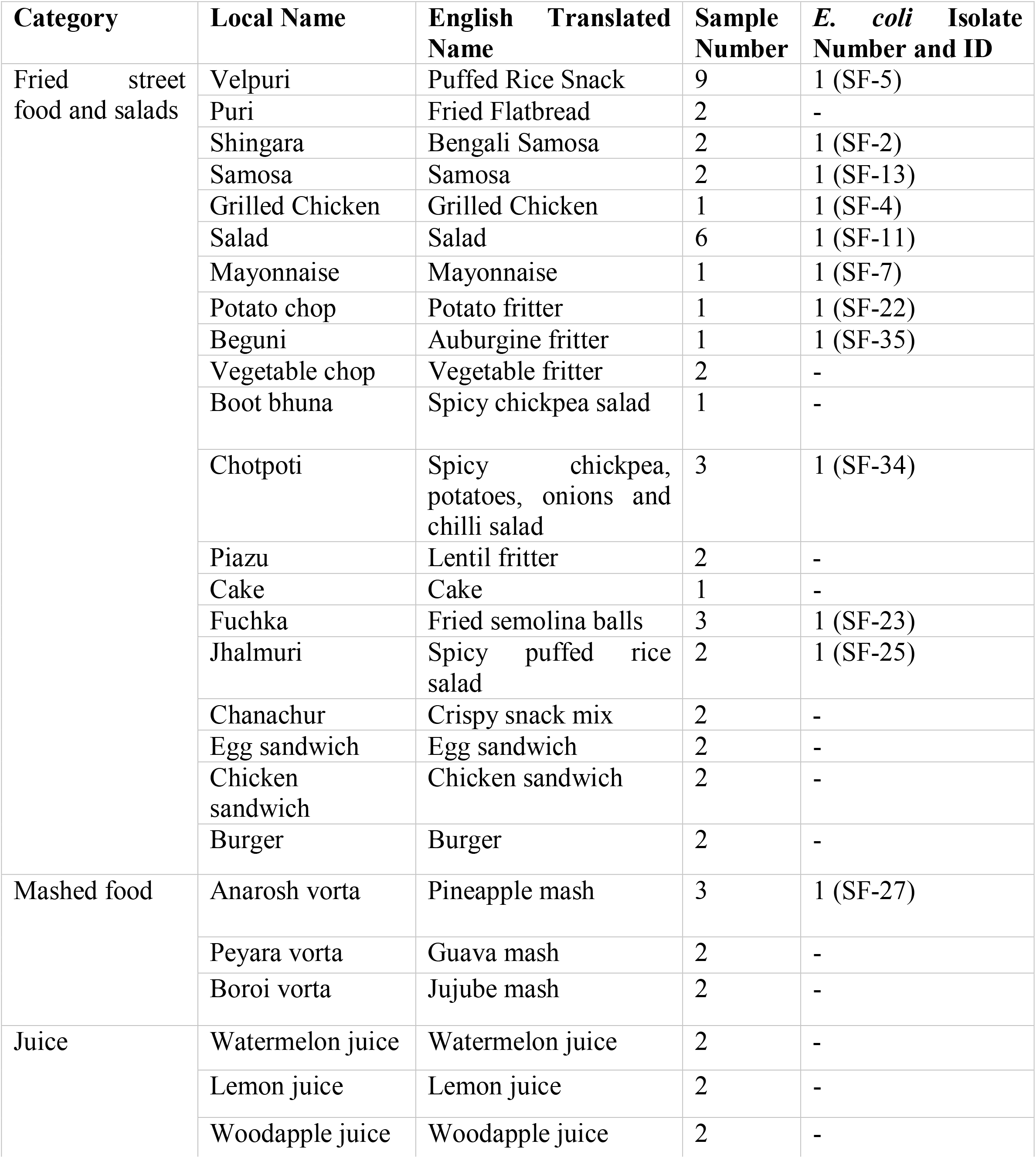
Food samples collected for this study.

After collection, samples were kept in labeled sterile polyethylene bags and transported in cool box containing ice blocks to the Environmental Microbiology Laboratory, Department of Microbiology, University of Dhaka, and immediately processed for bacteriological analysis.

### Sample Processing

For examination, 10 g of samples were weighed using electric balance and then blended using an automated blender machine for 2 minutes for homogenizing properly with 90 ml of phosphate-buffered saline (PBS) solution.

### Isolation and Primary Identification of *E. coli*

Coliform (including *E. coli*) counts were determined using the most probable number (MPN) method (Biswas et al., 2010). All presumptive and confirmatory tests of *E. coli* isolation as well as identification were performed according to Bhowmik et al. (2018).

### Gram staining and biochemical tests

Additional confirmation of test isolates was done by Gram staining and conventional biochemical tests (Bhowmik et al., 2019). Biochemical tests including Kligler’s Iron Agar (KIA) test, Citrate test, Motility, Urease (MIU) test, Methyl Red test as well as Voges–Proskauer test were performed for the identification of the isolates according to the methods described in the Microbiology Laboratory Manual (**C**appucino et al., 1996).

### Molecular confirmation of the presumptive *E. coli*

#### Template DNA

Colony PCR was carried out in every case of PCR reaction where the template DNA was prepared, as Mandal et al. (2014) said.

#### Master mix

PCR master mix contained deoxynucleotide triphosphates, Taq polymerase, MgCl_2,_ and buffer of 2X concentration (NEB, UK) (Mandal et al., 2014).

#### Identification of *E. coli* with Multiplex PCR

Multiplex PCR was conducted to detect two *E. coli* specific genes, *usp*A and *uid*A, for the reconfirmation of the tested isolates as *E. coli*. Primer sequences and PCR amplification reaction conditions were set up as described earlier (Godambe et al., 2017).

#### Amplified Ribosomal DNA Restriction Analysis (ARDRA)

The digestion patterns of the 16S rRNA gene PCR amplicons by restriction endonuclease enzymes from different species of bacteria may not be the same. Primers amplifying conserved sequences of the 16S rRNA gene were used as described earlier (Lu et al., 2000) for ARDRA. After purification, the PCR products were digested with three different restriction enzymes such as *Hae*III, *Hind*III, and EcoRI from New England BiolabsTM to more accurately identify *E. coli*. Initially, a master-mix was prepared with 7.25 μL autoclaved, filter-sterilized distilled water, 2 μL buffer, and 0.75 μL restriction enzyme solution. Then 10 μL of the master-mix was dispensed into 50 μL PCR tubes and 10 μL purified DNA was added to each of the tubes making up a total volume of 20 μL. The incubation period was 30 minutes at 37°C.

#### Phylogenetic grouping of *E. coli*

A quadruplex PCR previously described (Clermont et al., 2013) was conducted by targeting the marker *arp*A1 (400bp), *chu*A (288bp), *yja*A (211bp), and a DNA fragment, TspE4.C2 (152bp) to determine *E. coli* phylogroups (A, B1, B2, C, D, E, and F). Additionally, C-specific and E-specific PCR were performed for confirmation of phylogroups A/C, D/E, as well as E/Clade 1 by targeting the *trp*A (219 bp) and *arp*A2 (301bp) genes (Clermont et al., 2013).

#### Detection of *E. coli* virulence genes

According to conditions described previously, multiplex and singleplex PCR targeting different virulence genes (e.g., *eaeA, bfpA, elt, hlyA, ial*, and CVD432) were performed (Hegde et al., 2012).

#### Detection of class -1 integron

A singleplex PCR detected class 1 integron was conducted also following conditions described earlier (Mandal et al., 2014).

#### Plasmid profiling

The alkaline lysis method was used to investigate the presence of plasmids (Birnboim and Doly, 1979).

#### Resolution of PCR products

In this research 1.5% and 2% agarose gel electrophoresis were used to visualize all of the conventional PCR products and ARDRA, respectively. The sizes of PCR products were assessed using 100 bp DNA marker from GeneOn (UK) and GeneRuler (UK). Electrophoresis was carried out at 80-90 V for 1-2.5 hours, depending on the sizes of the amplicons.

In all cases, after electrophoresis, the gels were stained in ethidium bromide (40 µg/mL) for 30 minutes and then destained by running distilled water over the gels for 5 minutes. After that, the photographs were taken by a gel documentation system (Biorad, USA).

#### Antimicrobial sensitivity test

The antimicrobial susceptibility of the test isolates was conducted through the disc diffusion method. Muller-Hinton agar (Hi-media, India) media was used for this purpose. Isolates were tested against 13 commercially available antibiotics (Oxoid, UK), i.e., tetracycline (30 µg), ciprofloxacin (5 μg), ampicillin (10 µg), amoxicillin-clavulanic Acid (20 µg), nitrofurantoin (300 µg), azithromycin (15 µg), amikacin (30 µg), ceftriaxone (30 µg), cefixime (5 µg), chloramphenicol (30 μg), gentamycin (10 μg), imipenem (10 µg) and co-trimoxazole (25 µg). Test interpretation was made carefully according to the Clinical and Laboratory Standards Institute (CLSI) guideline, 2018 (Weinstein, 2018).

## Results And Discussion

### Presumptive identification of *E. coli* by Most Probable Number (MPN) method and plate-based assays

We collected 60 street food samples of different types from various sites in Dhaka city for this study. Among these 60 samples, 43 (71%) indicated the presence of lactose-fermenting and gas-producing bacteria in all three tubes of dilution series using inoculum quantities of 1.0, 0.1, and 0.01 g added to Brilliant Green Lactose Bile broth (BGLB). Therefore, we presumptively isolated a total of twelve (27% of samples, n=43)) isolates as *E. coli*. We then reconfirmed these isolates with different molecular tests, including PCR and ARDRA. Thus, the isolation rate of *E. coli* was similar to studies that we carried out earlier (Bhowmik et al., 2018; Bhowmik et al., 2019), where the rate of *E. coli* isolation from soil was 30.59%, and water was 26.88%.

### Identification of *E. coli* with PCR

We conducted a PCR assay to reconfirm twelve biochemically confirmed *E. coli*. We found each isolate to be positive for both targets in a multiplex PCR targeting two *E. coli* specific genes; *usp*A and *uid*A. (Figure 1). Through the same multiplex PCR technique, 149 isolates (79%) from food samples were identified as *E. coli* in a study by Godambe et al. (2017).

**Figure 1:**
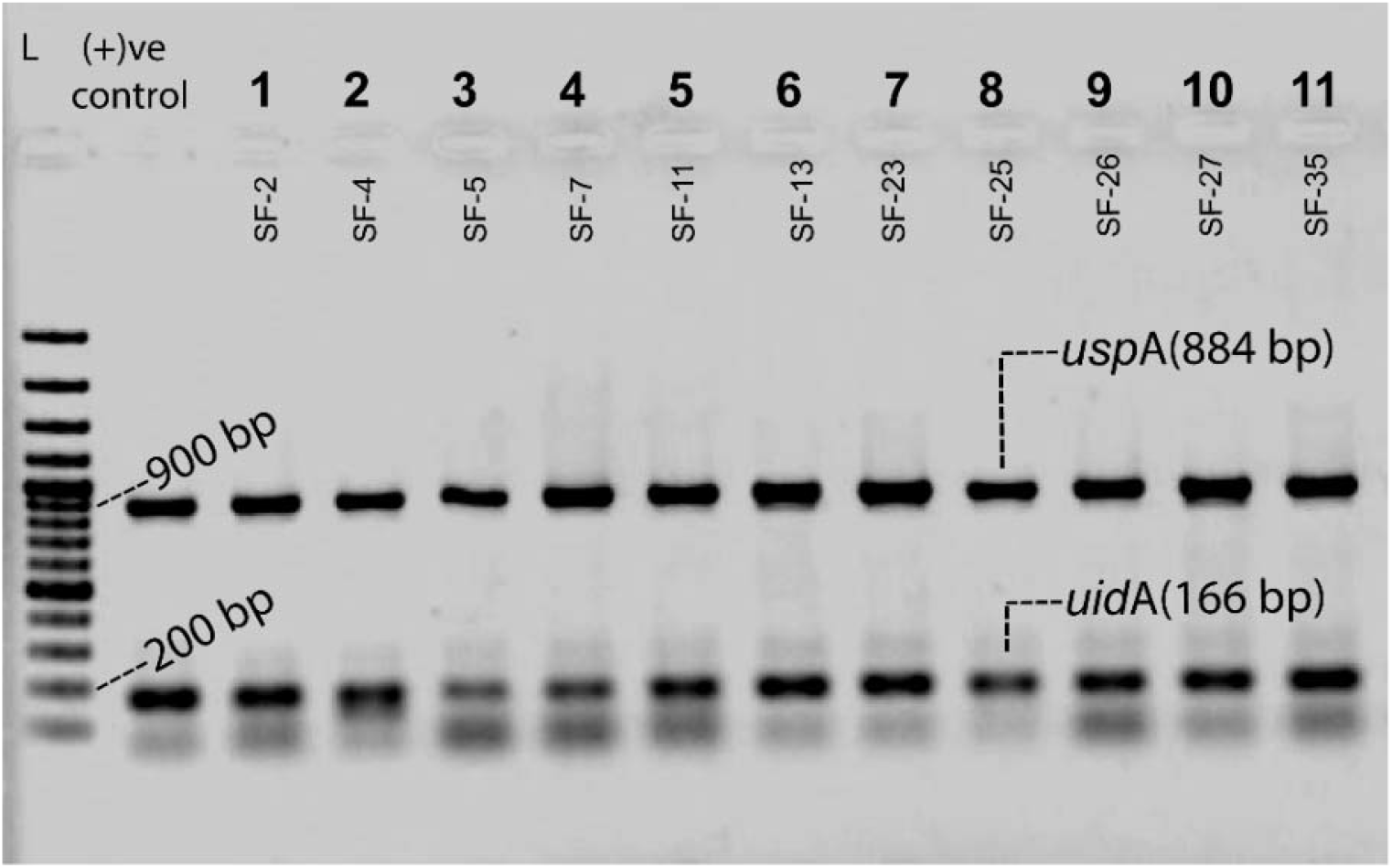
Agarose gel electrophoresis of PCR products for the detection of *E. coli* specific genes. In this study, we used *E. coli* ATCC 25922 as a positive control. The ladder was a 100 bp plus DNA ladder from GeneRuler (UK). All biochemically confirmed isolates were tested positive in this multiplex PCR.

### Typing of *E. coli* with ARDRA

ARDRA (Amplified Ribosomal DNA Restriction Analysis) was used to reconfirm the identification of 12 previously confirmed *E. coli* isolates. Purified 16s rRNA amplicons (Figure 2) were digested with three different restriction enzymes. Identical restriction fragment length patterns confirmed that the isolates were of the same type (Figure 3). Other studies have shown that ARDRA typing with restriction enzymes can be utilized to detect *E. coli* in clinical specimens (Lu et al., 2000).

**Figure 2.**
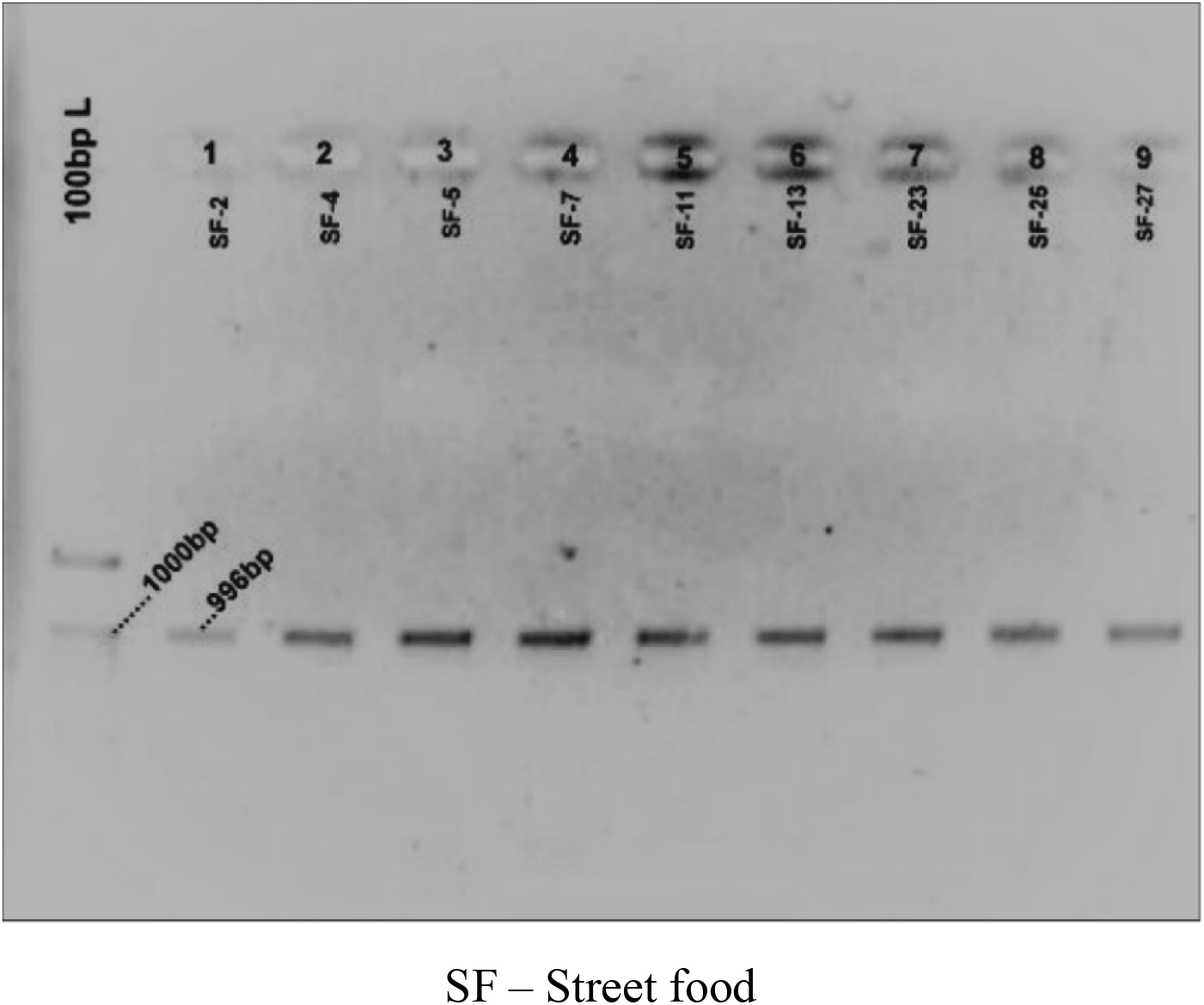
Agarose gel electrophoresis of 16S rRNA amplicon of *Escherichia coli*. Before moving into the ARDRA, purified amplicons of 16s rRNA PCR were visualized on 1.5% agarose gel for confirmation.

**Figure 3:**
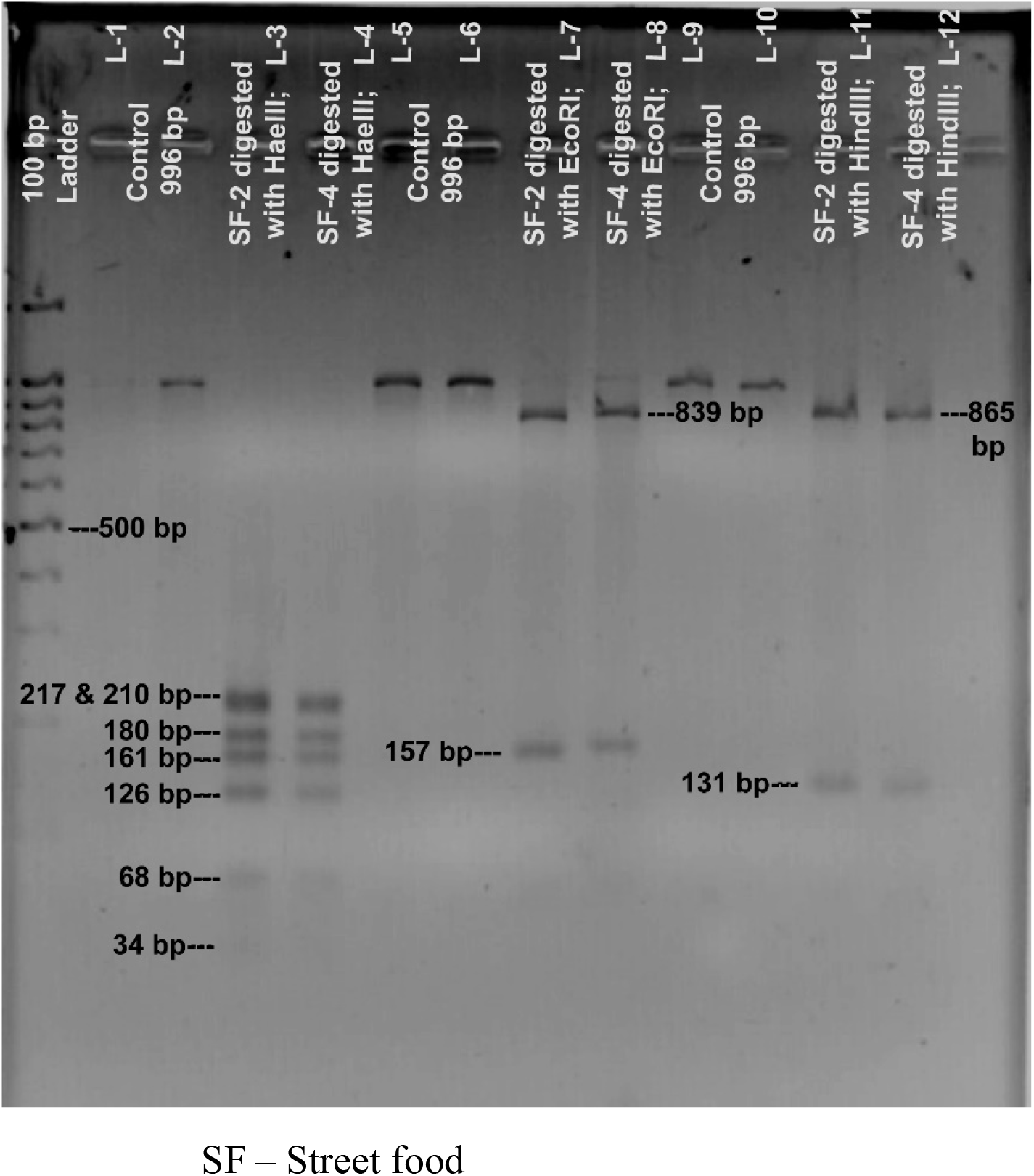
*Hae*III, *Eco*RI, *Hind*III digestion patterns of the purified 16S rRNA gene amplicons of few tested isolates. Samples in lanes 3 and 4 were *Hae*III digested PCR products from the control bacteria of lanes 1 and 2, labeled as L-1 and L-2 in this figure. Lanes 7 and 8 containing fragments of control DNA from L-5 and 6. Fragments in Lanes 11 and 12 were the *Hind*III - digested products of control DNA in L-9 and L-10. The ladder used was a 100bp Ladder from GeneON (UK).

### Prevalence of *E. coli* in food samples

In this research, *E. coli* was isolated at a rate of 8%. In a previous study (Hassan et al., 2016) a lower incidence of 3% (n=84) *E. coli* strains were found in the street food chotpoti **(3%**, n=84). Kharel et al. (2016) found *E. coli* in a variety of street food samples, including pakodas, and samosas. In a study conducted in Bangladesh, Khandoker et al. (2014) discovered *E. coli* and *Staphylococcus spp*. in spicy puffed rice (Jhalmuri). *E. coli* was shown in seafood, fruit juices, vegetable rice, and egg rice in India (Aruna et al., 2017). The authors declared that poor water quality and lack of hygiene practices during handling are responsible for food contamination. *E. coli* was found in 2.2% (n=60) of vegetable salad, cocoa drink, and ice kenkey in Ghana (Feglo and Sakyi, 2012). Untreated manure used as fertilizer for vegetable growth can become a reason behind *E. coli* contamination of vegetables like lettuce and tomatoes (Mensah et al., 2001). Previous research in Bangladesh (Rahman et al., 2017) found that *E. coli* was found in 37.86% (n=169) of foods of animal origin such as milk, chicken meat, and beef. In Kenya, *E. coli* has been identified on the street vended chicken, food contact surfaces, and equipment. Researchers concluded that the presence of flies and pests, dirty vending places, vending environments strewn with garbage, filthy clothes, and lack of hygienic knowledge might be responsible for *E. coli* contamination of such foods (Birgen et al., 2020). Sabuj et al. (2018), on the other hand, detected *E. coli* in shingara (20%, n=20), piazu (20%, n=15), potato chop (20%, n=20), beguni (20%, n=10), puri (20%, n=15), and samosa (20%, n=20). In Dhaka, Hossain et al. (2017) collected *E. coli* from street vendors selling velpuri, fuchka, salad, and water, much like we did. Furthermore, Biswas et al. (2010) identified the highest number of *E. coli* (90.90%, n=11) in vegetable-oriented ready-to-eat foods (RTE), rather than fish, meat, or cereal samples, in Bangladesh, and concluded that the presence of *E. coli* in RTE foods indicated possible fecal contamination. *E. coli* was also found in fruit juices (23%, n=86), milk shops (45% n=86), and dairy farms (45%, n=86) in Ethiopia, according to Tadesse et al. (2018). *E. coli* was found in velpuri (35%, n=20), masalapuri (30%, n=10), panipuri (30%, n=10), sevpuri (20%, n=5), and lime rice (20 percent, n=5), according to Bhaskar et al. (2004). Sample varieties, geographical locations, stress factors, sanitary management systems, and other factors all impacted the study’s outcomes (Mousa et al., 2020). In the present study, since street foods are mostly human handled in Bangladesh and vendors do not have enough concern about maintaining proper hygienic practices, we hypothesize that unclean hands’ improper handling led to *E. coli* in the samples studied.

### Distribution of phylogroups of *E. coli* in street foods

For identification of the origin of the 12 tested isolates, phylogroup PCR was conducted targeting different marker genes of *E. coli*. It was found that phylogroup B1 of environmental origin was the dominating group (58%), followed by the commensal group A (16%). Nonetheless, groups C, D, and E identified with a prevalence rate of 8% each. In a study conducted in Egypt, composed of 220 *E. coli*, derived from the dairy products by Ombarak et al. (2016), it was found that the phylogroup A (67.6%) was the predominant group followed by B1 (30.2%) and D (2.30%). Their finding conflicted with ours, and the larger sample number might become attributable to this discordance. However, in India, a study (Godambe et al., 2017) showed that the maximum number of *E. coli* (44%) among 192 *E. coli* isolates obtained from different food samples belonged to phylogroup B2, followed by group B1 (29%), A (16%) and group D (3%) in decreasing order. Their research also revealed that the virulent group B2 were isolated from different food samples of animal origin, including beef (72%), pork (53%), mutton (47%), and fish (33%), and pathogenic group D were isolated from chicken (18%) and sprouts (25%) (Godambe et al., 2017).

### Incidence of virulence genes in *E. coli*

The pathogenicity of *E. coli* depends mainly on the presence of different virulence genes and the existence of several toxins (Mousa et al., 2020). In the present study, out of 12 *E. coli* strains, only 8% from fuchka contained the virulence gene *hlyA* characteristic for EHEC and belonged to virulent phylogroup D (Figure 5). *E. coli* belonging to EHEC pathotypes can be responsible for bloody diarrhea and hemolytic uremic syndrome (HUS) (Karmali et al., 1983; Karmali, 1989; Griffin and Tauxe, 1991). Therefore, the identifying *hlyA* positive isolate from local street food is of particular concern. In an investigation in Egypt (Ombarak et al., 2016), it was found that out of 222 *E. coli* strains, 104 (46.8%) isolates from 69 different types of food samples carried one or more virulence genes where the prevalence of *hlyA* gene in *E. coli* identified from dairy products such as raw milk, Karish cheese, Ras cheese was 1.35%.

**Figure 4.**
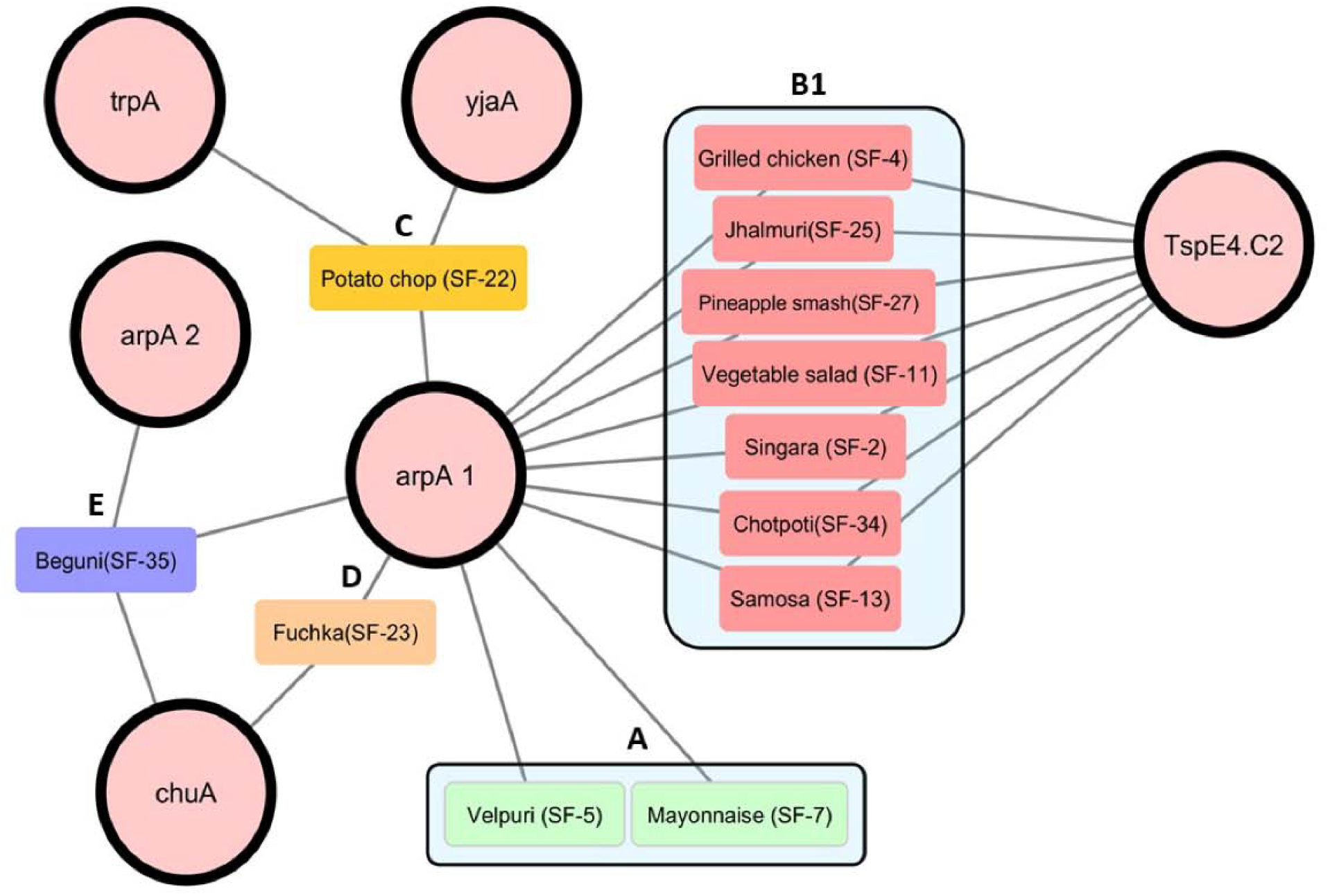
Graphical representation drawn using Cytoscape version 3.7.2 (Shannon et al., 2003) of the occurrence of genetic markers in *E. coli* strains isolated from different types of street foods. Large circles with black borders represent each genetic marker: *arp*A1, *chu*A, *yja*A, *trp*A, *arp*A2, and the DNA fragment TspE4.C2. Samples taken from different street foods are denoted by color-coded rounded rectangles and labeled accordingly. Edges between the genetic markers and a sample indicate that those genetic markers are detected in that sample. According to the presence/absence of specific genetic markers, the phylogroup of the samples was tagged, and black rounded rectangular borders highlight the phylogroup partitions.

**Figure 5.**
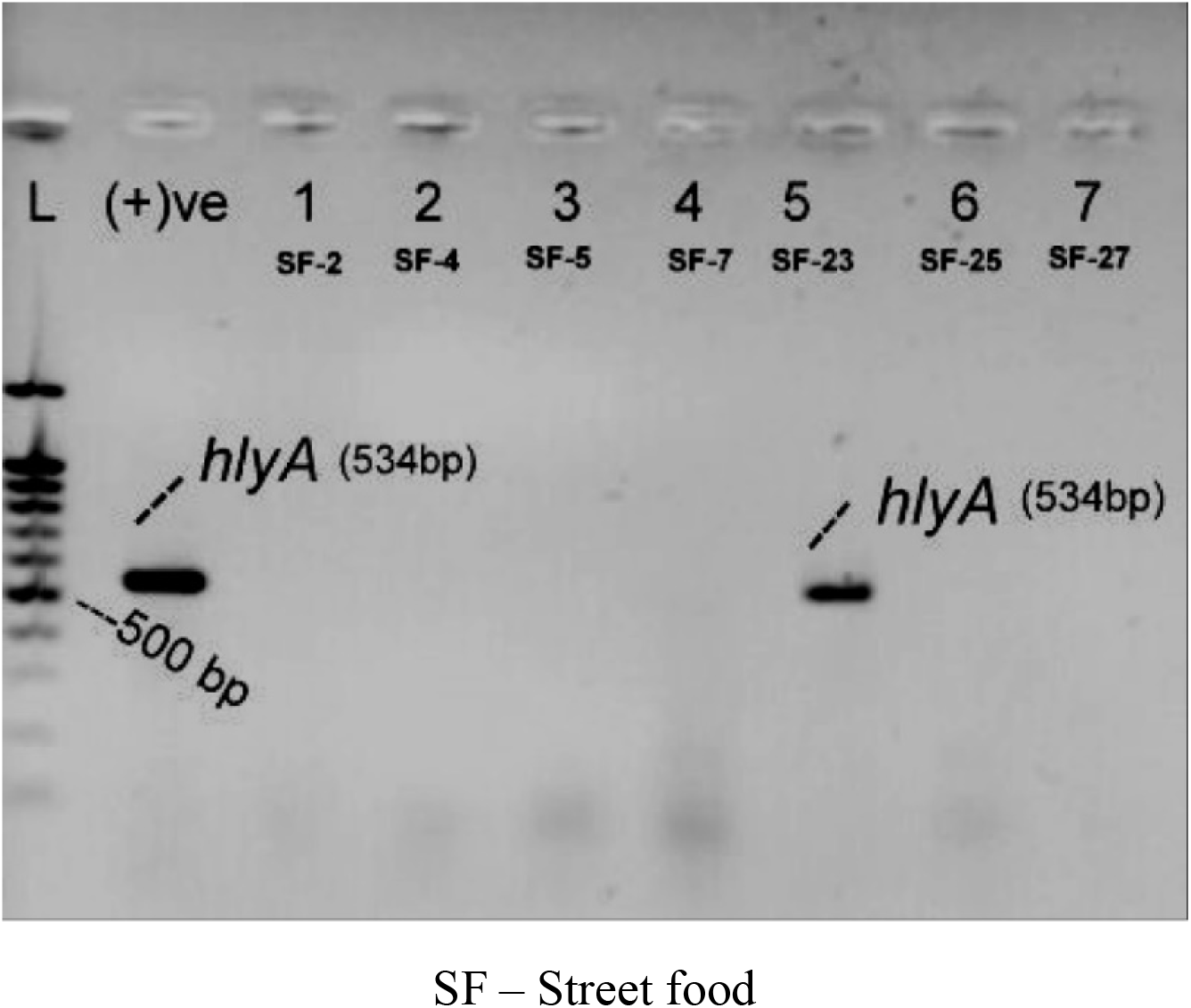
Detection of *hlyA* gene by agarose gel electrophoresis. The ladder, denoted as L is 100 bp ladder from GeneOn (UK). EHEC reference strain denoted as (+) ve was used as the positive control. Isolate in Lane 5 symbolized as SF-23 was positive for the *hlyA* gene.

### Antimicrobial susceptibility pattern of street food *E. coli*

In this study, the tested *E. coli* isolates showed the highest sensitivity to co-trimoxazole (83%) followed by chloramphenicol (75%). These study isolates were mostly resistant to ampicillin (100%), followed by azithromycin (66%), amikacin (66%), imipenem (58%), gentamycin (41%), and others (Figure 6 and Table 2). These are some of the most affordable and commonly available drugs for many developing countries like ours. This resistance percentage might be fatal gradually, leaving none of the treatment options available against infectious *E. coli*. A susceptibility pattern was observed for amoxicillin/clavulanic acid and cefixime (66% for each), where 58% of the isolates exhibited susceptibility against ceftriaxone and ciprofloxacin. The sensitivity pattern to other antibiotics such as tetracycline, gentamicin, azithromycin, imipenem, nitrofurantoin, and amikacin were 50%, 41%, 33%, 25%, 16%, and 8.33%, respectively. (Figure 6 and Table 2). Similar findings were reported by Sabuj et al. (2018), who showed that 100% (n=20) of their *E. coli* isolates from street food exhibited resistance to ampicillin, 80% against tetracycline, 30% against chloramphenicol and gentamicin (for each) followed by azithromycin (20%) but they observed that none of these isolates showed resistance against ciprofloxacin which is an exception to our finding. In a previous study (Hassan et al., 2017), sixteen *E. coli* isolated from street foods revealed the following sensitivity pattern: ciprofloxacin (76%), azithromycin (72%), sulfamethoxazole (84%), and gentamicin (56%), respectively. Their finding is almost similar to ours. Furthermore, another study on *E. coli* derived from fruit juice and raw milk (n=83) (Tadesse et al., 2018) reported that 70% of isolates showed resistance to ampicillin, similar to our findings. Additionally, they also reported that 100% of their isolates were sensitive to gentamicin followed by ciprofloxacin (90%) and tetracycline (60%), a finding similar to ours.

**Table 2.** Antibiogram profile of *E. coli* isolates from foods of different types

| Antimicrobial class | Antimicrobial agents | Disk codes and contents | No of *E. coli* isolates (%) |  |  |  |  |  |
| --- | --- | --- | --- | --- | --- | --- | --- | --- |
|  |  |  | R | % | I | % | S | % |
| Nitrofurantoin | Nitrofurantoin | F-300 | 3 | 25 | 7 | 58 | 2 | 16 |
| Beta-lactams | Amoxicillin/Clavulanic acid | AMC-30 | 2 | 16 | 2 | 16 | 8 | 66 |
| Macrolides | Azithromycin | AZM-30 | 8 | 66 | - | - | 4 | 33 |
| Cephalosporins | Ceftriaxone | CRO-30 | 2 | 16 | 3 | 25 | 7 | 58 |
| Chloramphenicol | Chloramphenicol | C-30 | 3 | 25 | - | - | 9 | 75 |
| Cephalosporins | Cefixime | CFM-5 | - | - | 4 | 33 | 8 | 66 |
| Aminoglycosides | Gentamicin | Gen-10 | 5 | 41 | 2 | 16 | 5 | 41 |
| Aminoglycosides | Amikacin | AK-30 | 8 | 66 | 3 | 25 | 1 | 8.33 |
| Fluoroquinolones | Ciprofloxacin | CIP-5 | 2 | 16 | 3 | 25 | 7 | 58 |
| Carbapenem | Imipenem | IPM-10 | 7 | 58 | 2 | 16 | 3 | 25 |
| Penicillin | Ampicillin | AMP-10 | 12 | 100 | - | - | - | - |
| Tetracycline | Tetracycline | TE-30 | 3 | 25 | 3 | 25 | 6 | 50 |
| Sulfonamides | Co-Trimoxazole | COT-25 | 2 | 16 | - | - | 10 | 83 |

**R-** Resistant, **I-** Intermediate Resistant, **S-** Sensitive

**Figure 6.**
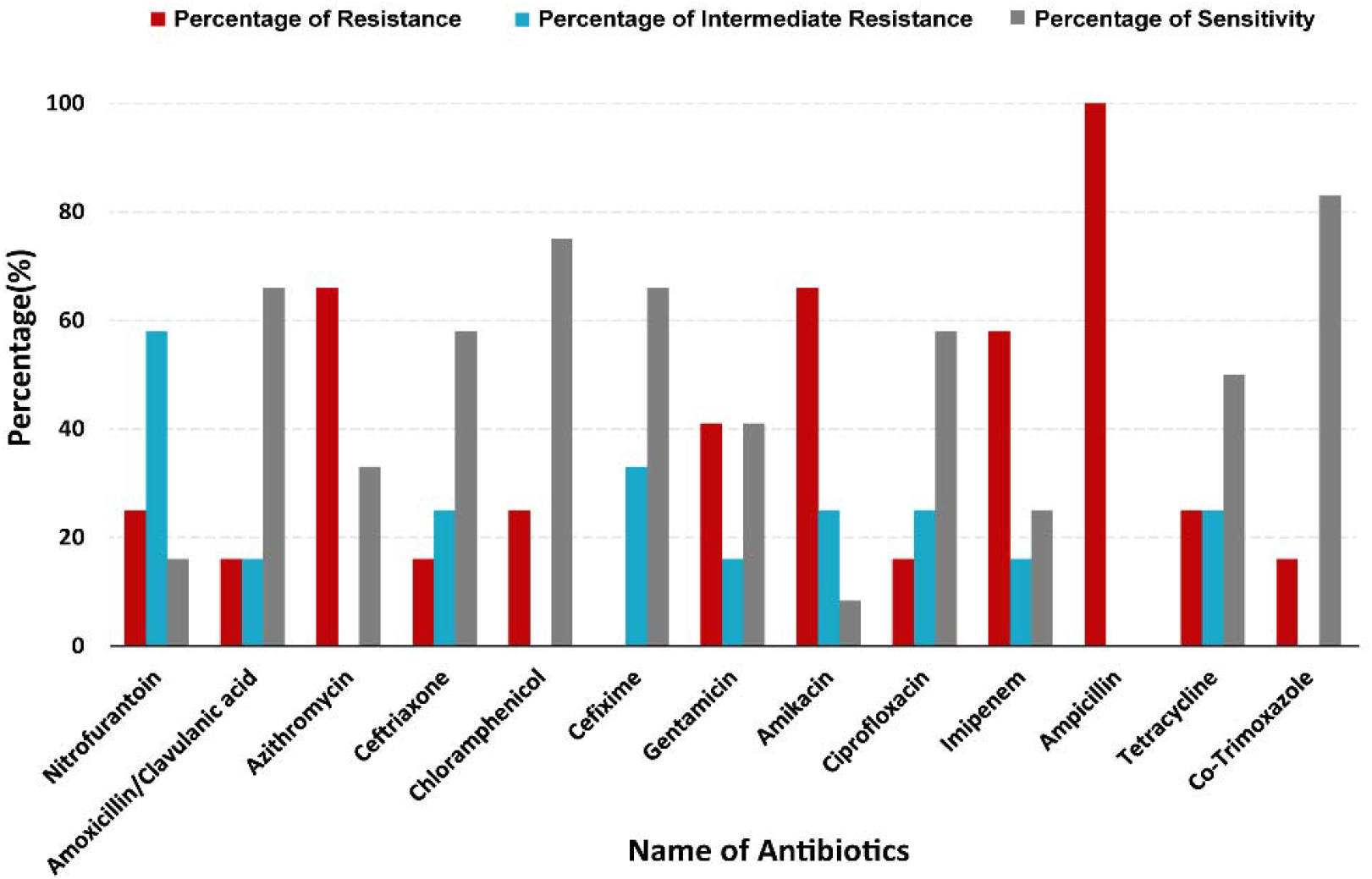
Antibiotic susceptibility profile of the study isolates to thirteen commercially available antibiotics.

**Figure 7.**
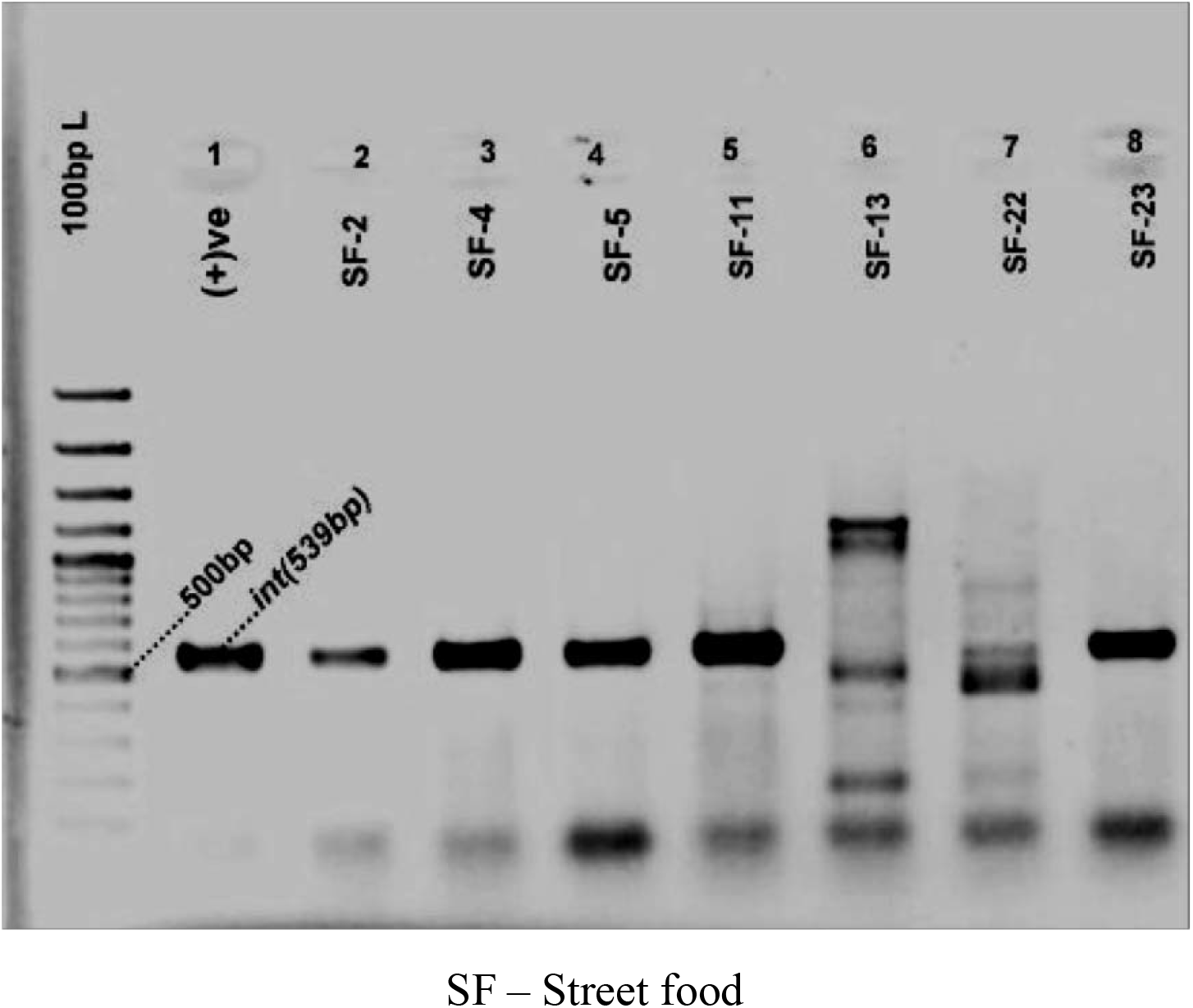
Agarose gel electrophoresis image of *intl-*1 PCR amplicon. Here, the ladder used was a 100 bp ladder from GeneON (UK). A laboratory reference *E. coli* known to contain *intl-*1 gene was used as the positive control in the PCR and denoted as (+)ve.

### Determination of the presence of class 1 integron and plasmid in the test isolates

The amplicon (539 bp) specific for class-1 integron was found in 41% of the 12 isolates; however, the plasmid was not detected in any of them. Pathogens from clinical settings, class 1 integrons are widely dispersed (Gillings et al., 2008). These are associated with the MDR seen in the hospital environment (Martinez-Freijo et al., 1998). Genes carried by integrons usually express multiple resistance mechanisms, such as resistance to beta-lactams, aminoglycosides, and other antimicrobial agents (Elbourne and Hall, 2006; Jeong et al., 2009). In the present research, it was noticed that class 1 integron did not correlate with multidrug resistance *E. coli* of street food.

## Conclusion

The findings of this study led us to believe that street foods are potential reservoirs of *E. coli* from the environment (phylogroup B1) and, to a lesser extent, of commensal *E. coli* (phylogroup A). All of these isolates showed resistance against most of the tested antibiotics. They also carried class 1 integrons. This indicates that multidrug-resistant *E. coli* can act as potential reservoirs of antibiotic resistance in street foods. Proper hygiene and sanitation conditions must be maintained during the preparation, handling, and delivery of such foods to avoid contamination with *E. coli*.

## Data Availability

All data produced in the present study are available upon reasonable request to the authors

## Acknowledgement

The author would like to express thanks to the Department of Microbiology, University of Dhaka for laboratory facilities.

